# Trends in hospitalizations and mortality rates for anti-neutrophil cytoplasmic antibody-associated vasculitis in Mexico: Results from a Nationwide Health Registry

**DOI:** 10.1101/2025.03.13.25323860

**Authors:** Ivet Etchegaray-Morales, Claudia Mendoza-Pinto, Susana Barrera-Hernández, Yolanda Martina Martínez-Barragán, Pamela Munguía-Realpozo, Ingris Pelaez-Ballestas, Roberto Berra-Romani, Edith Ramírez-Lara, Jorge Ayón-Aguilar, Socorro Méndez-Martínez

## Abstract

Anti-neutrophil cytoplasmic antibody (ANCA)-associated vasculitis entails substantial morbidity and mortality, yet no epidemiologic evidence exists on its outcomes in Mexico. This study assessed national hospitalizations (2005–2022) and mortality (2000–2022) related to ANCA-associated vasculitis using data from the General Board of Health Information. We extracted hospital discharges and deaths classified under ICD-10 M31.3 and M31.7, computing age-standardized hospitalization (ASHR) and mortality (ASMR) rates per 100,000 persons, overall and by sex. Annual percentage change (APC) and average annual percent change (AAPC) were estimated with Joinpoint regression. We identified 2,804 hospitalizations and 599 deaths. Females accounted for 50.3% of hospitalizations, while males represented 48.7% of deaths. Although overall ASHR and ASMR AAPCs were not statistically significant, notable trends emerged. From 2010 to 2022, ASHR declined significantly (APC: −5.2%; 95% CI: −9.7, −0.5; p=0.03), whereas mortality rates remained stable from 2000 to 2022 (AAPC: +3%; 95% CI: −4.6, 11.3; p=0.45). Nevertheless, mortality increased among males (APC: +6.4%; 95% CI: 0.9, 12.2; p=0.02) and individuals over 45 years (APC: +8.6%; 95% CI: 1.7, 16.0; p=0.02) from 2008 onward. Overall, these findings indicate no major changes in national rates, but reveal a decline in hospitalizations since 2010 and a rise in mortality for specific subgroups since 2008. Targeted interventions, particularly for older adults and men, appear warranted to address this evolving disease burden. Future research should explore underlying risk factors and evaluate tailored strategies to improve clinical outcomes in ANCA-associated vasculitis across Mexico. These insights may guide policy decisions and contribute to improved patient management nationwide.

## Introduction

Granulomatosis with polyangiitis (GPA) and microscopic polyangiitis (MPA) are systemic antineutrophil cytoplasmic autoantibodies (ANCA)-associated vasculitis (AAV), which represent chronic and recurrent autoimmune diseases marked by small-vessel necrotizing inflammation and circulating ANCA in the majority of patients (60–90%) at disease onset [1, 2].

Although the etiology and pathogenesis of AAV are multifaceted, genetic factors play a role that partially accounts for the observed geographical differences [3, 4]. Clinically, GPA and MPA include the respiratory system and glomerulonephritis (GN), with less frequent neurologic and cutaneous involvement. Both GPA and MPA are primarily treated with glucocorticoids (GCT) with rituximab or cyclophosphamide for severe conditions and azathioprine or methotrexate for moderate [5]. Despite some progress in diagnosis and management, morbidity and mortality remain relevant [6].

Patients with GPA and MPA can have frequent hospitalizations, and previous studies have shown a mortality rate of 30% to 43% among hospitalized patients with AAV [7]. According to a systematic review, mortality is approximately 3-fold higher among AAV patients than the general population [8]. Additionally, global patterns in overall mortality of AAV found that in 2014, the age-standardized mortality rate (ASMR) was 0.53 deaths per million inhabitants, slightly more pronounced in males than females [9]. The same study identified that in Latin America, after initial stability between 2001 and 2005, the ASMR significantly increased until 2014. In contrast, the mortality trends in other regions showed either a decrease or stability. Although the causes of those trend differences are unclear, a previous study showed that Hispanics with AAV exhibit more severe disease and elevated damage indices [10]. However, a recent study observed a remarkably low mortality rate due to vasculitis in Hispanic populations versus non-Hispanic people [11]. The researchers hypothesized that mortality rate discrepancies may be explained by underdiagnosis of vasculitis or distinct genetic susceptibilities compared to other ethnic groups. Currently, epidemiological studies on ANCA-associated vasculitis mortality and hospitalization in Mexico are lacking. Therefore, we aimed to evaluate the variation in Mexico’s ANCA-associated vasculitis hospitalizations and mortality during the 2005-2022 and 2000-2022, respectively.

## Methods

### Data sources and case definition

A nationwide retrospective data analysis was performed using mortality and hospital discharge records from the dynamic information cubes recorded by the General Directorate of Health Information (DGIS) Open Access datasets belonging to the Mexican Health Ministry. These databases compile annual hospital discharges and mortality statistics and systematically collect records from all Mexican states and public hospitals, ensuring comprehensive nationwide coverage [12]. Inherent to methodological issues, only cases coded as ANCA-associated vasculitis as the leading cause of hospital discharge or death were included.

Hospitalization records extracted from the DGIS Open Access datasets do not contain individual-level identifiers, so it was impossible to distinguish whether multiple hospitalizations originated from the same patient. Therefore, each hospitalization was considered as an independent event in our analysis.

Different study periods were used to ensure robust analyses based on data availability. Hospitalization data were available from 2005 to 2022, while mortality data were analyzed from 2000 to 2022 due to the earlier availability of mortality records in the DGIS system. This distinction allows for an accurate assessment of trends while utilizing the most comprehensive datasets available.

This study was approved by the Local Research Ethics Committee of the Instituto Mexicano del Seguro Social (IMSS) (R-2023-2106-011), and due to its retrospective nature, the requirement to obtain informed consent from the patients was waived.

Available ANCA-associated vasculitis registers in subjects aged 15 and over for hospital discharges and mortality in 2005-2022 and 2000-2022, respectively, using the M31.3 and M31.7 codes of the International Classification of Diseases (ICD), 10^th^ revision. Data from the National Institute of Statistics, Geography, and Informatics (INEGI, by its Spanish acronym) for each year during the same period (2005-2022 for hospital discharge and 2000-2022 for deaths) were used to measure mortality in the general population (total and subgroup-specific) [13]. For each record, information on sex, age, and geographic region was collected. Geographic region was categorized according to the INEGI classification (Northern, Central, Midwest, and Southeast) (Supplementary Table 1).

## Statistical analysis

To standardize yearly comparisons of hospitalization and mortality rates, we used the estimated 2010 population distribution to calculate the overall age-standardized hospitalization rate ASHR and ASMR for ANCA-associated and non-ANCA-associated vasculitis, as described previously [14]. To ensure consistency in trend analysis, we used the estimated 2010 population distribution as the reference for calculating ASHR and ASMR. The decision to use 2010 rather than a midpoint of the study period was based on several methodological considerations. First, the 2010 census provides an official, stable, and well-characterized population structure from the INEGI, ensuring consistency with national health surveillance practices. Second, using 2010 minimizes potential biases from demographic shifts over time, as census-based population estimates offer greater accuracy than interpolated projections for non-census years. Finally, this approach aligns with previous epidemiological studies in Mexico, facilitating comparability with national and international literature [14].

We then computed the annual ASHR and ASMR values of people with ANCA-associated vasculitis and the non-ANCA vasculitis population. We summarized trends by sex and age groups (15–19, 20–29, 30–39, 40–49, 50–59, 60–69, 70–79, and ≥80 years). Time trends of direct age-standardized rates for ANCA-associated vasculitis were analyzed using the Joinpoint regression analysis [15]. We calculated the annual percent change (APC) and 95% confidence intervals (CIs) by applying a linear regression to the natural logarithm of the age-standardized rates, with the calendar year as an independent variable. The average APC (AAPC) described the average APCs over the study periods.

In addition, a subgroup analysis was conducted based on sex (female and male), age group (15-44 and 45 years or more) and geographic region. However, due to the potential for inaccuracies in mortality rate calculations, subgroups with ≤20 deaths were excluded from the analysis.

The Joinpoint Regression Program (v5.2.0.0) from the US National Cancer Institute Surveillance Research Program was used (http://surveillance.cancer.gov/joinpoint) [15]. A significance threshold of α = 0.05 was used for the test.

## Results

### General characteristics of ANCA-associated vasculitis hospitalizations and deaths

ANCA-associated vasculitis was recorded as the underlying cause of hospitalizations and deaths in 2,804 (GPA: 2671 and MPO: 133) and 599 (GPA: 546 and MPO: 53) patients in Mexico from 2005 through 2022 and from 2000 through 2022, respectively. The mean of hospital stay lasted 8.5 ± 10.7 days. The proportions of hospitalizations and deaths by sex were similar (Table 1). The proportion of deaths in those aged ≤ 44 and those aged was higher for ANCA-vasculitis than for non-ANCA-associated vasculitis deaths (48.4% vs. 15.2%, respectively; *p* < 0.001). On the other hand, the proportion of hospitalizations for the same age group was lower for ANCA-associated vasculitis than non-ANCA-associated vasculitis (48.4% vs. 66.7%, respectively; *p* < 0.001).

**Table 1.** Number and average annual rates for ANCA-vasculitis hospitalizations (2005-2022) and mortality (2000-2022) in Mexico.

| **Characteristics** | **Hospitalizations**<br>**n (%)** | **AAPC of hospitalizations rate**<br>**per 100 000 population (95% CI)** | **Deaths n**<br>**(%)** | **AAPC of mortality rate per**<br>**100 000 population (95% CI)** |
| --- | --- | --- | --- | --- |
| **Overall** | 2,804 (100) | -1.5 % (-15.8, 15.3) | 599 (100) | 3.0 % (-4.6, 11.3) |
| **Sex** |  |  |  |  |
| Male | 1,410 (50.3) | -0.2 % (-7.7, 7.9) | 292 (48.7) | 0.5 % (-4.9, 6.2) |
| Female | 1,394 (49.7) | -0.8 % (-17.9, 19.9) | 307 (51.3) | 7.3 % (-7.5, 24.5) |
| **Age group**<br>**(years)** |  |  |  |  |
| 15-44 | 1,356 (48.4) | 0.1 % (-6.6, 7.2) | 221 (36.9) | 3.5 % (-8.5, 16.2) |
| 45 or more | 1,448 (51.6) | -0.9 % (95 % CI: -4.9, 3.2) | 378 (63.1) | 2.5 % (-4.3, 9.7) |
| **Region** |  |  |  |  |
| Northern | 412 (14.7) | 2.59 % (95% CI: -12.5, 20.4) | 122 (20.4) | 0.9 % (-24.7, 35.4) |
| Midwest | 504 (17.9) | 5.2 % (-8.2, 20.6) | 148 (24.7) | 16.2 % (7.3, 25.9) |
| Central region | 1,732 (61.8) | 1.3 % (-18.5, 25.8) | 286 (47.7) | 5.2 % (-1.4, 12.2) |
| Southern | 156 (5.6) | 4.7 % (-23.2, 42.8) | 43 (7.2) | NA* |

**AAPC: average annual percent change; CI: confidence interval; NA: not applicable.**
**\*NA: the calculated age-adjusted rate is zero.**

### Overall hospitalizations and mortality rates trends in ANCA and non-ANCA associated vasculitis populations

Overall, the ASHR decreased without statistical significance from 2005 through 2022 (AAPC: -1.5 %; 95% CI: -15.8 to 15.3; *p* = 0.36) (Table 1), whereas this rate of non-significant increase in the non-ANCA-vasculitis population (AAPC 2.27 %; 95% CI: -5.90 to 11.17; *p* = 0.59). The segmented analysis observed a significant rate decrease between 2010 and 2022, with an APC of -5.19% (95% CI: -9.71, - 0.45, p = 0.03) for ANCA-vasculitis patients.

Although the ASMR during the study period was stable (Table 1), the joinpoint analysis for the mortality rate identified segmented variations over the study period (Table 2). From 2000 to 2008, the mortality rate exhibited a minimal increase with an APC of 0.6% (95% CI: -6.8% to 8.5%, p = 0.88). A notable shift occurred between 2008 and 2018, where mortality rates significantly increased with an APC of 7.8% (95% CI: 1.1% to 14.9%, p = 0.03). However, from 2018 to 2022, the trend reversed, showing a decrease in mortality rates with an APC of -8.6% (95% CI: - 26.7% to 14.1%, p = 0.40). In contrast, the non-ANCA-associated vasculitis group did not exhibit significant variations over time (AAPC: 0.70%; 95%CI: -0.34 to 1.77, *p* = 0.18). A statistically significant uptrend was identified in males from 2008 through 2016 (APC: 16.2%; 95% CI 7.3 to 25.9; *p =* 0.001), whereas no variations were found in females.

**Table 2.** The APC of ANCA-vasculitis hospitalization rates among Mexican aged ≥15 years.

| Populatio<br>n | Period 1 |  |  | Period 2 |  |  | Period 3 |  |  |
| --- | --- | --- | --- | --- | --- | --- | --- | --- | --- |
|  | Years | APC (95% CI) | P | Years | APC (95% CI) | P | Years | APC (95% CI) | P |
| **Overall** | 2005-2007 | -12.7(-61,99.1) | 0.72 | 2007-2010 | 24.7 (-45.3,<br>184.7) | 0.56 | **2010-**<br>**2022** | **-5.2% (-9.7, -**<br>**0.5)** | **0.03** |
| **Female** | 2005-2007 | -19.9(-70.5,<br>117.4) | 0.63 | 2007-2010 | 27.6(-<br>52.9,246.5) | 0.59 | 2010-2022 | -3.4 (-9, 2.3) | 0.20 |
| **Male** | **2005-**<br>**2012** | **10.1% (0.3,**<br>**20.9)** | **0.04** | 2012-2017 | -15.2(32.1,5.7) | 0.12 | 2017-2022 | 2.5 (-12.3, 19.8) | 0.73 |
| **15-44**<br>**years** | 2005-2012 | 12.3 (-2.6,<br>29.5) | 0.90 | 2012-2015 | -22.5 (-73.4,<br>125.5) | 0.61 | 2015-2022 | 2.4 (-11.3, 1.8) | 0.10 |
| **45 or more**<br>**years** | 2005-2014 | 4.9 (-1.2, 11.5) | 0.10 | 2014-2020 | -8.8 (-21.3,<br>5.6) | 0.19 | 2020-2022 | 2.0 (-47.2, 97.2) | 0.95 |
| **Northern** | 2005-2012 | 4.6(-13,25.8) | 0.59 | 2013-2016 | -23.2(-<br>85.8,316.1) | 0.73 | 2016-2022 | 23.2 (-7.3, 64.0) | 0.13 |
| **Midwest** | **2005 -** | **5.6% (-**<br>**10.6, -0.4)** | **0.03** | 2014-2018 | 32.1(-<br>26.3,137) | 0.31 | 2018-2022 | -0.9 (-31.5,<br>43.3) | 0.95 |

| **2014** |  |  |  |  |  |  |  |  |  |
| --- | --- | --- | --- | --- | --- | --- | --- | --- | --- |
| **Central** | 2005-2015 | -3.2(-11.4,5.7) | 0.42 | 2015-2018 | 38.7(-<br>55.7,334.8) | 0.53 | 2018-2022 | -18.2(-43,17.2) | 0.24 |
| **Southern** | 2005-2015 | -3.5(-11.6,5.3) | 0.38 | 2015-2018 | 68.8(-<br>45.5,423.2) | 0.32 | 2018-2022 | -14.9(-<br>40.5,21.6) | 0.33 |

**Abbreviations: AAPC= average of annual percentage change; APC= Annual percentage change.**

**Table 3.**
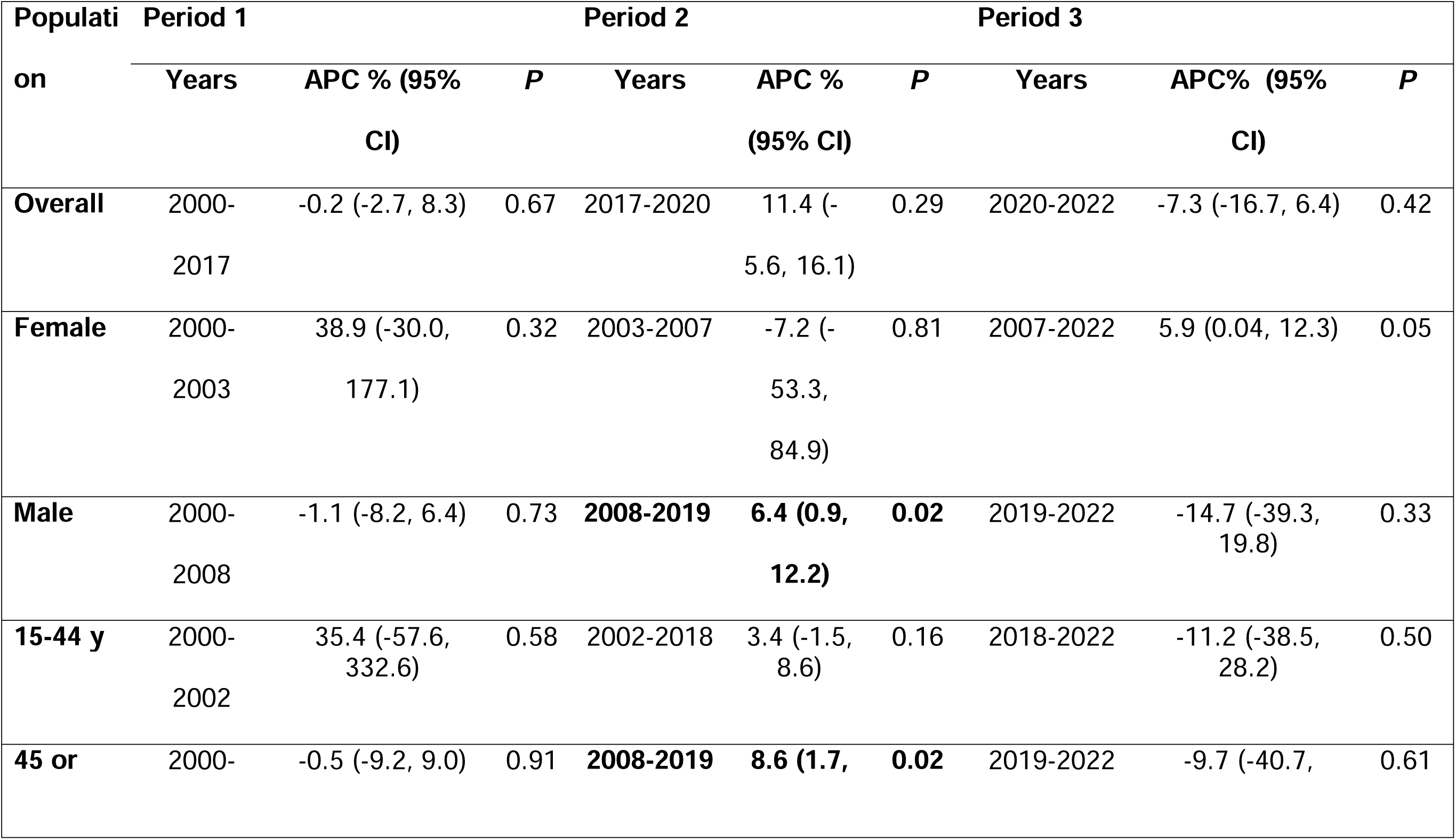
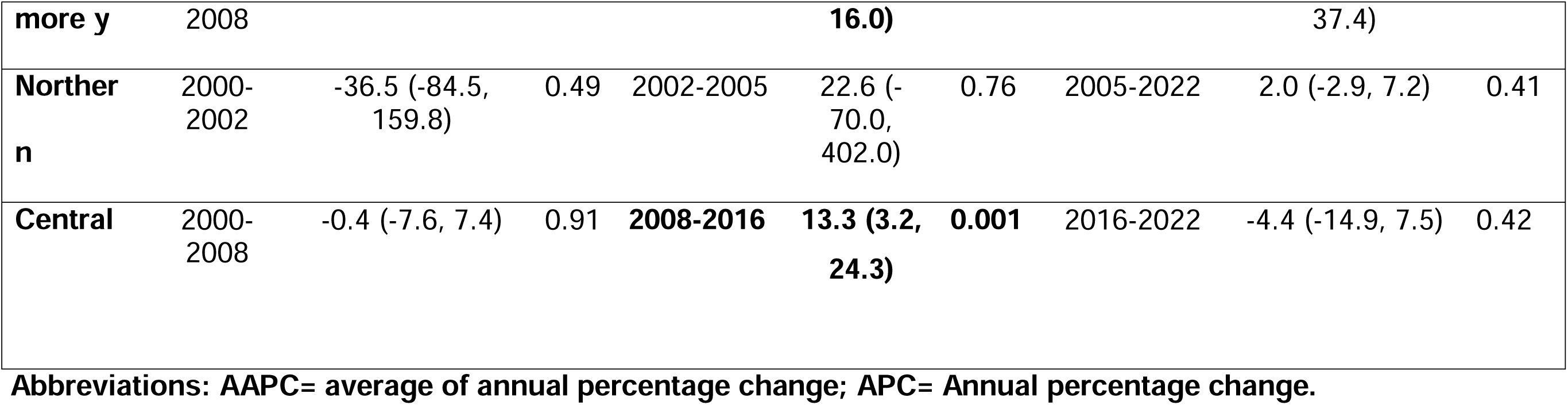
The APC of ANCA-vasculitis mortality rates among Mexicans aged ≥15 years.

### Trends in hospitalizations and mortality rates by sex and age group in ANCA-associated vasculitis population

Hospitalization rates in men increased significantly between 2005 and 2012, with an APC of 10.1% (95% CI: 0.3 to 20.9, p = 0.04) (Table 2). A notable increase in ASMR was recorded in men between 2008 and 2019, with an APC of 6.4% (95% CI: 0.9 to 12.2, p = 0.02) for ANCA-associated vasculitis patients. However, no significant changes were identified in women. A significant increase was identified in the group over 45 years of age between 2008 and 2019, with an APC of 8.8% (Fig 1).

**Figure 1.**
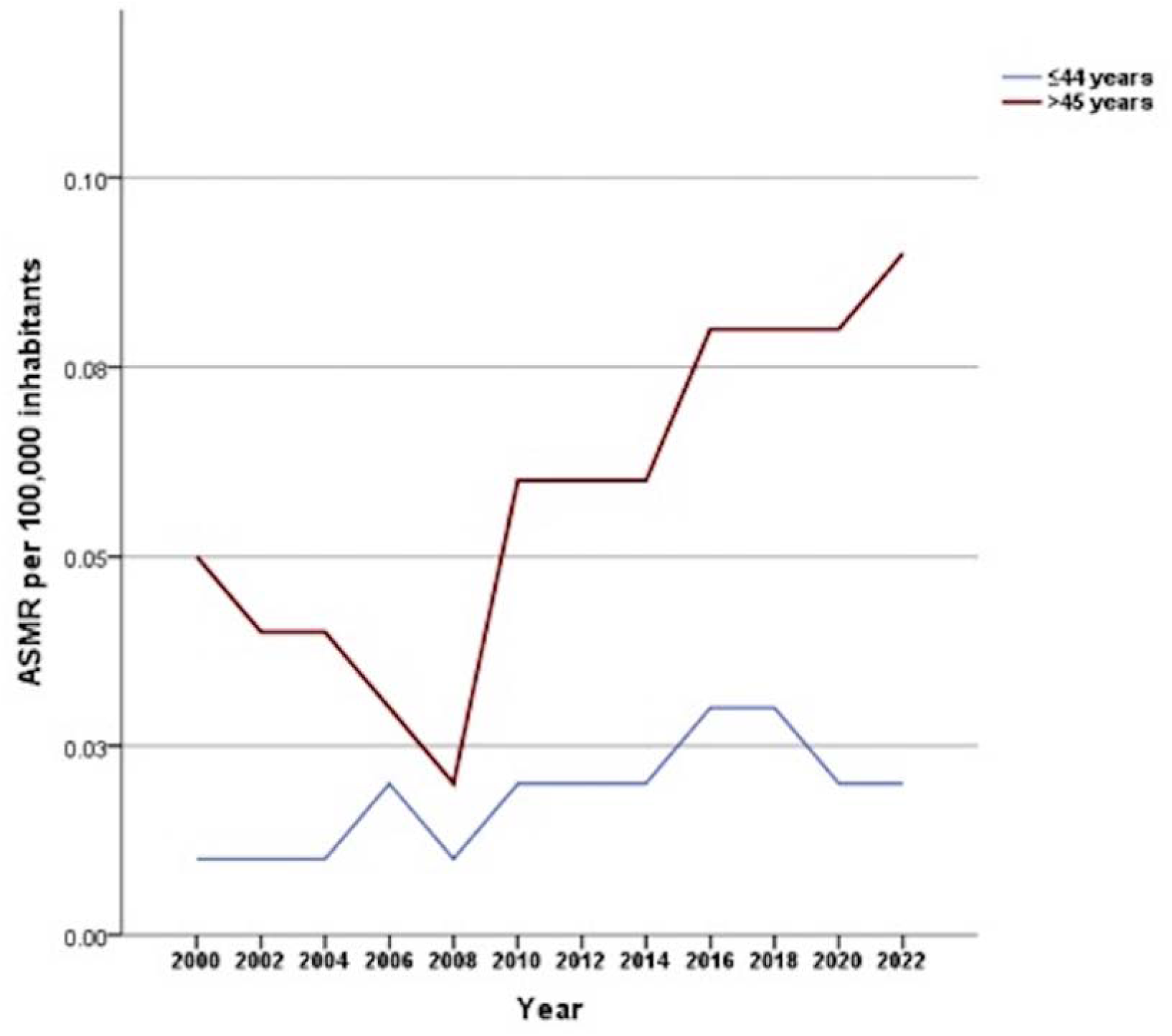
Age-standardized mortality rates (ASMR) for ANCA-associated vasculitis in Mexico from 2000 to 2022, stratified by age group.

### Trends in hospitalizations and mortality rates by geographic regions in ANCA-associated vasculitis population

Regarding the analysis by geographic regions, the hospitalization rates in all four areas increased overall; however, the AAPCs were not significant for the North, Midwest, Central, and Southeast regions, respectively (Table 1). Specifically in the Midwest region, a significant decrease was detected during the study period from 2005 to 2014, with an APC of -5.6% (95% CI: -10.6 to -0.4, *p* = 0.03).

In the North and Central regions, mortality rates increased, though not significantly, with an AAPC of 0.9% (95% CI: -24.7 to 35.4, p = 0.95) and 5.2% (95% CI: -1.4 to 12.2, *p* = 0.12), respectively. In the Central region, rates were increased between 2008 and 2016, with an APC of 13.3% (95% CI: 3.2 to 24.3, *p =* 0.001). Due to the low frequency of deaths in the Southeast and Midwest regions, a trend analysis could not be performed.

Although the AAPCs for ASMR by geographic regions were not statistically significant over time, the Central region showed a statistically significant uptrend (APC: 16.2%; 95% CI: 7.3 to 25.9; *p =* 0.001).

## Discussion

The results indicate a significant decrease in age-standardized hospitalization rates from 2010 to 2022, reflecting an overall downward trend in hospital discharges for ANCA-associated vasculitis. In contrast, the overall ASMR remained relatively stable across the study period. Additionally, the findings highlight sex-specific, age-group, and geographic regional disparities, with males, older, and certain geographic regions, particularly the Central region, experiencing higher increases in mortality. These results emphasize the importance of focused interventions to reduce these inequities and advance health outcomes in populations affected by ANCA-associated vasculitis in Mexico.

The significant decrease in ASHR for ANCA-associated vasculitis observed between 2010 and 2022 likely reflects multiple factors, including advances in the early diagnosis and management of the disease, which were also implemented in Mexico. One critical development is refining ANCA-associated vasculitis classification criteria, particularly the 2012 Revised International Chapel Hill Consensus Conference nomenclature [2] and subsequent updates [16]. These have improved the ability to diagnose and categorize ANCA-associated vasculitis subtypes accurately. These criteria have enhanced diagnostic precision [17], leading to earlier identification of patients with ANCA-associated vasculitis and potentially allowing for more effective outpatient management, thereby reducing hospitalizations. Additionally, advancements in diagnostic technologies, such as the increased availability of ANCA testing, have facilitated the early detection of the disease [18, 19]. Coupled with more effective immunosuppressive therapies, such as rituximab and cyclophosphamide, and improved glucocorticoid-sparing regimens, these developments have improved disease control, preventing severe exacerbations and hospital admissions [20, 21]. Public health initiatives to increase awareness of autoimmune diseases may have also encouraged earlier medical intervention [22–24]. However, this trend must be interpreted in light of healthcare accessibility, as socioeconomic disparities in Mexico might still affect the reduction in hospitalizations, with some populations potentially underrepresented due to challenges in accessing specialized care [25, 26]. Further investigation is required to determine whether the observed decrement in age-standardized hospitalization rate reflects actual improvements in disease management or if persistent gaps influence healthcare accessibility.

Our findings of increased mortality trends for ANCA-associated vasculitis in Mexico from 2008 to 2018 stand in contrast to the global decrease in AAV mortality reported by Scherlinger et al. [9], who observed a significant annual decline of 1.01% in the worldwide age-standardized mortality rate (ASMR) from 2001 to 2014. While their study highlighted geographic regional differences, with Europe showing higher ASMRs than Asia and Latin America, our findings suggest a distinctive trend in Mexico, where mortality has risen in recent years, particularly among older individuals, males, and Central region residents. In addition, a French nationwide study by Bataille et al. [27] revealed a stable but elevated standardized mortality ratio (SMR) for ANCA-associated vasculitis over an eight-year period (2010–2017). The SMRs for GPA and MPA were consistent, ranging from 2.03 to 2.66, indicating a two- to three-fold higher mortality risk than the general population. Notably, five-year survival rates differed significantly between the two conditions, with GPA patients demonstrating higher survival rates (81%) than those with MPA (72%), reflecting variations in disease severity and comorbidities. These findings are consistent with our study, which also identified persistently high mortality rates among AAV patients despite therapeutic advances.

The marked increase in mortality rates associated with ANCA-associated vasculitis from 2008 to 2019 in some population groups reflects an evolving burden that warrants further investigation. Although our study did not directly evaluate factors underlying this trend, several potential explanations emerge. This increase may be attributed to factors such as heightened disease severity, delayed diagnosis, and complications arising from treatment [28–30]. Additionally, the widespread use of immunosuppressive therapies may have rendered patients more susceptible to infections, cardiovascular complications, and other comorbidities, contributing to the observed increase in mortality [30, 31]. These potential factors emphasize the need for future studies to directly assess underlying drivers of mortality in ANCA-associated vasculitis to inform targeted interventions and improve patient outcomes in our country.

The findings from this study underline notable sex-specific, age-group, and regional disparities in mortality associated with ANCA-associated vasculitis in Mexico, with older males and residents of the Central region showing disproportionately higher increases in mortality rates. Older populations may bear a disproportionate burden of AAV-related mortality, likely influenced by greater comorbidity prevalence, delayed diagnosis, or differences in treatment responses. The differential burden by geographic region suggests that local healthcare access, socioeconomic factors, and regional variations in disease management may contribute to these disparities [32–34]. This concentration of mortality in the Central region may partly reflect Mexico City’s role as a healthcare hub, where specialized hospitals and medical centers are more accessible [34, 35]. Given the rarity of ANCA-associated vasculitis, patients from surrounding areas likely seek care in Mexico City, leading to a higher recorded mortality rate due to the aggregation of more severe or complex cases. Furthermore, geographic disparities might reflect environmental or genetic factors that could influence disease susceptibility and severity, particularly in diverse populations such as those in Mexico. The observed higher mortality rates in males align with broader literature [36–38], indicating that men with ANCA-associated vasculitis may experience more severe disease manifestations, higher rates of comorbid conditions such as cardiovascular disease, and potentially different treatment responses. This study’s identification of such disparities highlights the need for targeted public health strategies and resource allocation to regions and demographics experiencing higher mortality risks, emphasizing the importance of equitable access to specialized care and tailored interventions in managing ANCA-associated vasculitis effectively across Mexico.

The study period (2000–2022 for mortality and 2005–2022 for hospitalizations) includes the COVID-19 pandemic (2020–2021), significantly affecting healthcare utilization patterns and mortality rates worldwide. However, recent evidence from Mexico suggests that the observed mortality rates for systemic autoimmune rheumatic diseases (SARD), including ANCA-associated vasculitis [39]. Several factors, including potential underreporting, changes in healthcare access, and altered disease management patterns during the pandemic may explain the lack of a substantial increase in mortality.

While offering valuable insights into hospitalization and mortality trends for ANCA-associated vasculitis in Mexico, this study has several limitations. First, a retrospective analysis based on registry data is subject to potential coding inaccuracies, particularly concerning the cause of death and primary diagnosis, which may lead to misclassification bias. Additionally, the reliance on aggregate data from national health registries limits the ability to assess individual-level clinical characteristics, such as disease severity, specific comorbidities, or treatment regimens, which could significantly influence outcomes. Furthermore, as patients with severe disease may be more likely to seek care in specialized hospitals concentrated in urban areas like Mexico City, regional mortality rates may be inflated due to the clustering of severe cases, thus limiting the generalizability of findings to rural or underserved regions. Another limitation is the lack of data on socioeconomic factors and healthcare access, which may play a critical role in understanding regional and sex-specific disparities in mortality. A notable limitation of this study is the inability to analyze trends specifically for GPA and MPA separately due to mainly the low number of hospitalizations and deaths attributed to these subtypes in the dataset, which restricted the statistical power needed to detect meaningful trends over time. In addition, a key limitation of our study is the inability to distinguish between first-time hospitalizations and readmissions, as the dataset does not contain individual-level identifiers. Consequently, hospitalizations were analyzed as independent events, possibly leading to overestimating hospitalization rates if some patients were admitted multiple times. However, this approach remains a standard methodology in epidemiological studies based on administrative health registries, particularly when assessing national-level hospitalization burden trends over extended periods. Lastly, because ANCA-associated vasculitis is a rare disease, the limited sample size over the study period may reduce the statistical power to detect subtle trends and associations. Given these limitations, there is a clear need for prospective studies that include thorough clinical, demographic, and socioeconomic data to provide a more nuanced understanding of the factors driving mortality trends in AAV in Mexico.

In summary, this research offers a crucial understanding of the hospitalization and mortality trends associated with ANCA-associated vasculitis in Mexico, revealing a decrease in hospitalization rates from 2010 to 2022, possibly reflecting advances in disease management and early diagnosis. However, the observed increase in mortality from 2008 to 2018 highlights an evolving burden, likely influenced by factors such as disease severity, comorbidity prevalence, or treatment-related complications, although these factors were not directly assessed in this study.

Regional and sex-specific disparities, particularly the higher mortality in males and the Central region, highlight the role of healthcare infrastructure, as Mexico City’s concentration of specialized facilities may attract complex cases from across the country. These findings call for targeted public health strategies to improve equitable access to specialized care, address comorbidity risks, and enhance disease management practices across all geographic regions. Future prospective studies that incorporate individual-level clinical and socioeconomic data are essential for understanding and addressing the drivers of mortality in AAV, ultimately working towards reducing mortality and improving outcomes for patients with this rare autoimmune disease in Mexico.

## Data Availability

All data produced in the present study are available upon reasonable request to the authors

