## Supplementary material for "Trends in hospitalizations and mortality rates for anti-neutrophil cytoplasmic antibody-associated vasculitis in Mexico: Results from a Nationwide Health Registry"

**Table S1.** Geographic regions, according to The National Institute of Statistics and Geography (INEGI)

**Northern**

Baja California, Baja California Sur, Chihuahua, Coahuila, Durango, Nuevo León, Sinaloa, Sonora and Tamaulipas

**Midwest**

Aguascalientes, Colima, Guanajuato, Jalisco, Michoacán de Ocampo, Nayarit, Querétaro, San Luis Potosí and Zacatecas

**Central Region**

Guerrero, Hidalgo, Mexico City, State of Mexico, Morelos, Puebla, Tlaxcala and Oaxaca

**Southern**

Campeche, Chiapas, Quintana Roo, Tabasco, Veracruz de Ignacio de la Llave and Yucatán
